# Increased circulating cell-free DNA in eosinophilic granulomatosis with polyangiitis: implications for eosinophil extracellular traps and immunothrombosis

**DOI:** 10.1101/2021.08.04.21261617

**Authors:** Teppei Hashimoto, Shigeharu Ueki, Yosuke Kamide, Yui Miyabe, Mineyo Fukuchi, Yuichi Yokoyama, Tetsuya Furukawa, Naoto Azuma, Nobuyuki Oka, Hiroki Takeuchi, Kyoko Kanno, Akemi Ishida-Yamamoto, Masami Taniguchi, Akira Hashiramoto, Kiyoshi Matsui

**Author notes:** **Corresponding author:** Teppei Hashimoto, Division of Diabetes, Endocrinology and Clinical Immunology, Department of Internal Medicine, Hyogo College of Medicine, 1-1, Mukogawa, Nishinomiya 663-8501, Japan.

## Abstract

**Background:** Endogenous DNA derived from nuclei or mitochondria is released into the blood circulation as cell-free DNA (cfDNA) following cell damage or death. cfDNA is associated with various pathological conditions; however, its clinical significance in antineutrophil cytoplasmic antibody-associated vasculitis (AAV) remains unclear. This study aimed to evaluate the clinical significance of cfDNA in AAV.

**Methods:** We enrolled 35 patients with AAV, including 10 with eosinophilic granulomatosis with polyangiitis (EGPA), 13 with microscopic polyangiitis, and 12 with granulomatosis with polyangiitis. Serum cf-nuclear DNA (cf-nDNA) and cf-mitochondrial DNA (cf-mtDNA) levels were measured by quantitative polymerase chain reaction. Tissue samples from EGPA patients were examined by immunofluorescence and transmission electron microscopy. The structure, stability, and platelet adhesion of eosinophil extracellular traps (EETs) were also assessed *in vitro*.

**Results:** Serum cf-nDNA and cf-mtDNA levels were significantly higher in AAV than in healthy controls, with the highest levels in EGPA; however, serum DNase activities were comparable among all groups. cf-nDNA and cf-mtDNA decreased after treatment and were associated with disease activity only in EGPA. Blood eosinophil count and plasma D-dimer levels were significantly correlated with cf-nDNA in EGPA and cf-mtDNA. EGPA tissue samples showed lytic eosinophils and EETs in small-vessel thrombi. EETs showed greater stability against DNase than neutrophil extracellular traps and provided a scaffold for platelet adhesion *in vitro.*

**Conclusion:** cfDNA was increased in EGPA, associated with disease activity. The presence of DNase-resistant EETs might contribute to the occurrence of immunothrombosis in EGPA.

## Introduction

Endogenous DNA derived from nuclei or mitochondria is released into the blood circulation as result of the damage or death of peripheral blood cells and tissues. This extracellular DNA, referred to as cell-free DNA (cfDNA), is thought to derive primarily from dead cells of the hematopoietic lineage, with minimal contributions from other tissues^1,2^. Levels of nuclear-derived cfDNA (cf-nDNA) are significantly increased in cancer patients and can be used to monitor disease activity^3-5^, while lower concentrations of cf-nDNA are present in peripheral blood from healthy individuals^6^. Mitochondria-derived cfDNA (cf-mtDNA) has also been reported as a promising diagnostic and prognostic biomarker in several types of cancer^7^. Recent studies have indicated the potential roles of cf-nDNA and cf-mtDNA in autoimmune diseases, especially rheumatoid arthritis and systemic lupus erythematosus (SLE)^8-10^. cfDNA is mainly cleared by DNase1, and DNase deficiency thus leads to the persistence of DNA/chromatin complexes in the serum, leading to the development of autoimmune diseases^11^. Low serum DNase1 activity has been reported in patients with SLE and has been associated with elevated cfDNA levels^12,13^.

Antineutrophil cytoplasmic autoantibody (ANCA)-associated vasculitis (AAV) is characterized by pauci-immune vasculitis and the presence of ANCA. AAV includes granulomatosis with polyangiitis (GPA), microscopic polyangiitis (MPA), and eosinophilic granulomatosis with polyangiitis (EGPA)^14^. AAV is a life- or organ-threatening disease treated with high-dose glucocorticoids and immunosuppressants^15^. Over 90% of patients with GPA and MPA have ANCA autoantibodies against proteinase 3 (PR3) or myeloperoxidase (MPO), which are related to the pathological mechanism of the diseases^16^. ANCA can activate neutrophils to release inflammatory cytokines and induce cell death, referred to as NETosis, which in turn releases filamentous DNA coated with histones, i.e. neutrophil extracellular traps (NETs). NETs contain granular and cytoplasmic proteins, including the targeted autoantigens PR3 and MPO^17^. Excess formation of NETs can be pathogenic, causing thrombosis and vascular endothelial cell injury^18^. EGPA is differentiated from GPA and MPA by features associated with eosinophilia and asthma, with 30%–40% of patients with these conditions being ACNA-positive^19^. Eosinophil extracellular traps (EETs) have also been associated with allergic diseases^20-22^, although the pathological role of EETs in EGPA is less well understood.

Previous studies reported increased concentrations of cfDNA in the serum of patients with GPA^23^ and with remission EGPA^24^. However, the relationships between different types of AAV and cfDNA remain unclear. The primary aim of this study was to evaluate the clinical significance of cfDNA in patients with AAV.

## Materials and methods

### Study subjects

The study included 35 patients with newly diagnosed AAV at Hyogo College of Medicine. Peripheral blood samples were collected before and after immunosuppressive therapy. The patients were diagnosed according to the methodology for the classification of AAV^25^. Patients with EGPA fulfilled the 1990 American College of Rheumatology criteria^26^. Serum samples were also collected from 10 healthy age- and sex-matched controls (HC) (62.6 ±15.4; 3 male, 7 female). Biopsy samples (2 skin and 7 sural nerve tissue) from nine patients with newly-diagnosed EGPA were obtained before immunosuppressive treatment. Peripheral blood was obtained from healthy donors for experimental protocols requiring purified eosinophils and neutrophils. The study was designed and conducted according to the Helsinki declaration and approved by the institutional ethics committees of Hyogo College of Medicine (approval no. 0374) and Akita University Graduate School of Medicine (approval no. 994). Written informed consent was obtained from all patients. Disease activity was monitored according to the Birmingham vasculitis activity score version 3 (BVAS)^27^.

### Preparation and quantification of cfDNA

Blood samples were separated to produce serum and cfDNA was extracted using a Qiamp MinElute ccfDNA kit (Qiagen, Hilden, Germany). The concentration of cf-nDNA was measured by quantitative polymerase chain reaction (qPCR) for ALU repeats, using a LightCycler 480 Instrument II (Roche Diagnostics, Basel, Switzerland), as described previously^28^. Long and short DNA fragments were detected using primers amplifying the 115-bp amplicon (ALU115) of total cf-nDNA. The sequences of the ALU primers were as follows: forward primer 5′-CCTGAGGTCAGGAGTTCGAG-3′ and reverse primer 5′-CCCGAGTAGCTGGATTACA-3′. The concentration of cf-mtDNA was also measured by qPCR with primers amplifying the 79-bp fragments of the mitochondrial 16s-RNA gene, with the short fragment representing total mtDNA^29^. The 79-bp mtDNA primer sequences were as follows: forward primer 5′-CAGCCGCTATTAAAGGTTCG-3′ and reverse primer 5′-CCTGGATTACTCCGGTCTGA-3′. The detailed qPCR procedure is shown in Supporting Materials and Methods.

### Measurement of DNase1 activity and eosinophilic cationic protein (ECP)

Details are provided in Supporting Materials and Methods.

### Immunofluorescence staining

Tissue samples were fixed with 10% formalin and embedded in paraffin for pathological tissue analyses. For major basic protein (MBP) and galectin-10 staining, deparaffinised sections underwent antigen retrieval with 0.1% protein kinase K at room temperature (RT) for 6 minutes, followed by blocking with phosphate-buffered saline (PBS) containing 10% bovine serum albumin (BSA). The sections were then incubated with primary rabbit anti-human MBP antibody (10 μg/ml; kind gift from Dr. Hirohito Kita, Mayo Clinic, Scottsdale, AZ, USA) for 30 minutes at 37°C and mouse anti-galectin-10 antibody (B-F42; 1:50 dilution; Cat. No. ab27417, Abcam, Cambridge, MA, USA) for 90 minutes at RT. The sections were then incubated with Alexa-488-conjugated goat anti-mouse IgG antibody (1:200 dilution; Cat. No. A11001, Life Technologies, Carlsbad, CA, USA), Alexa-594 goat anti-rabbit IgG antibody (1:200 dilution, Cat. No. A11072, Life Technologies), and Hoechst 33342 (1:5000 dilution; Cat. No. H3570, Invitrogen, Carlsbad, CA, USA) for 30 minutes at RT.

For citrullinated histone H3 (CitH3) staining, sections underwent antigen retrieval by incubation in Tris-ethylenediaminetetraacetic acid buffer in a microwave oven for 15 minutes. Sections were then blocked with PBS containing 10% BSA and incubated with primary rabbit anti-CitH3 monoclonal antibody (GR194277-3, 10 μg/ml, Abcam) and mouse anti-CD31 antibody (187377, 1 μg/ml, Abcam) for 90 minutes at RT, followed by Alexa-488–conjugated goat anti-rabbit antibodies (1:200 dilution, Cat. No. A11008, Life Technologies), Alexa-594 goat anti-mouse IgG antibody (1:200 dilution, Cat. No. A11005, Life Technologies), and Hoechst 33342 for 30 minutes at RT. Isotype-matched control antibodies were used in each experiment. Samples were mounted using Prolong Diamond (Life Technologies) and images were obtained using a Carl Zeiss LSM 780 confocal microscope (Carl Zeiss, Oberkohen, Germany). In some experiments, coverslips were removed and the sections were further stained with hematoxylin-eosin (H-&E) after observation of the immunofluorescent images. Immunostaining and H&E images were compared to confirm that all cells and tissues were damage-free.

### Electron microscopy

For transmission electron microscopy (TEM), sural nerve tissues from EGPA patients were fixed and prepared as described previously^30^. Scanning electron microscopy (SEM) was used for isolated eosinophils and neutrophils^21^. The details are described in Supporting Materials and Methods.

### Cell preparation

Eosinophils were isolated from peripheral blood using a MACS™ system (Miltenyi Biotec, Bergisch Gladbach, Germany) with CD16-negative selection (anti-CD16 antibody-conjugated microbeads, #130-045-701, Miltenyi Biotec) as described previously^31^. The purity of the isolated eosinophils was >98% of nucleated cells and the cell viability was >99%. Neutrophils (>95% neutrophils, viability >98%) were obtained using the same system. Platelet-rich plasma was obtained by centrifugation of citrated blood at 150 × *g* for 10 minutes.

### Quantification of extracellular trap (ET) area

Isolated eosinophils and neutrophils (1 × 10^5^) were stimulated for 180 minutes at 37°C in 0.3% BSA/RPMI medium with 10 ng/ml phorbol 12-myristate 13-acetate (PMA), as a potent inducer of EETs and NETs^21^. No mitocondrial DNA traps^22^ were observed in our experimental settings (**Supporting video**). The ET area was quantified by adding SYTOX green (1:5000, #S7020, Life Technologies) to the medium. Fluorescent images were obtained and viewed under an inverted microscope (DMI 4000B, Leica, Tokyo, Japan) equipped with a Cooled Color Digital camera (DFC450C, Leica). Three randomly selected images were processed in 8-bit black/white with manual thresholding to obtain binary images using ImageJ software, and the numbers of pixels were measured. There were at least 30 cells per image. The SYTOX area per cell was then calculated.

### DNase-induced ET dissolution assay

To induce ETosis, isolated eosinophils and neutrophils (2 × 10^5^/well in clear-bottomed 96-well plate) were stimulated overnight with PMA in 0.3% BSA RPMI, resulting in cell death in100% of cells and at ETosis in >95% of cells^21^. Double-stranded DNA (dsDNA) was quantitated by adding Picogreen (Thermo Fisher Scientific, Waltham, MA USA) and shaken at 200 rpm using an MS1 plate shaker (IKA Works, Wilmington, NC, USA), followed by excitation at 485 nm and measurement of fluorescence at 520 nm using a fluorescence microplate reader (BMG Labtech, Ortenberg, Germany). DNase1 (10 ng/ml, final concentration) or vehicle control was added to each well and the fluorescence intensity was measured at the indicated time points. The ratio of fluorescence intensity between DNase1 and control (DNase1/control × 100) at 0 minutes was considered as baseline.

### Platelet adhesion assay

Isolated eosinophils (2 × 10^6^/ml in µ-slide IV, ibidi, Gräfelfing, Germany) were stimulated overnight with PMA in 0.3% BSA RPMI to induce EETosis. The medium was replaced with Ca^2+^-containing Hanks balanced salt solution and the slides were incubated with or without DNase1 (40 U/ml) for 10 minutes at 37°C. The platelet-rich plasma concentration was adjusted to 3 × 10^8^/ml using an automated hematology analyzer (Sysmex XE-5000, Sysmex, Kobe, Japan) and recalcified by adding 1 mM CaCl_2_. Platelets were labeled with fluorescent calcein-AM (1 µM, Invitrogen) and loaded in µ-slide, followed by incubation for 60 minutes at 37°C. After gentle washing the slide gently with HBSS, random images were obtained using a LSM 780 confocal microscope (20x objective; Carl Zeiss). Platelets were counted in a coded manner. For SEM sample preparation, a non-labeled platelet suspension was similarly incubated on the slide for 60 minutes at 37°C.

### Statistical analysis

Statistical analysis was carried out using JMP Pro version 14.2 (SAS Institute Inc. Cary, NC, USA). Comparisons between two groups with equal variance were performed using two-sided Student’s *t*-tests. Differences between continuous variables were analyzed by Mann–Whitney U tests and differences between categorical variables were analyzed using Fisher’s exact tests. Comparisons among multiple groups were analyzed by Kruskal–Wallis tests. Correlations between cfDNA and BVAS and serological parameters were determined using Spearman’s rank correlation coefficient. A difference of *P*<0.05 was considered significant.

## Results

### Patient characteristics

The clinical characteristics of immunosuppressive therapy-naive patients with AAV are summarized in **Table 1**. The neutrophil count was significantly higher in patients with MPA compared with those with EGPA or GPA, and the eosinophil count and serum ECP were significantly higher in patients with EGPA. There were no significant differences among the groups in terms of age, C-reactive protein, and BVAS. Two patients with EGPA, two with GPA, and one with MPA had thrombosis (deep vein thrombosis). The types of medication and the clinical characteristics of the patients when cfDNA was measured after immunosuppressive therapy are also shown in **Table S1**. Patients with EGPA were initially treated with high-dose glucocorticoids and immunosuppressants based on the 2009 5-factor score (FFS)^32^. In this study, when FFS ≥1 and initial glucocorticoid therapy had insufficient effect, rituximab was added. Other patients were treated with glucocorticoids with or without azathioprine. Patients with GPA or MPA and organ-threatening disease received high-dose glucocorticoids plus rituximab or cyclophosphamide. The duration of glucocorticoid therapy and the types of immunosuppressants used were comparable among all groups. The BVAS score improved after immunosuppressive therapy in all groups.

**Table 1.** Patient characteristics at baseline

|  | EGPA (n=10) | MPA(n=13) | GPA(n=12) | p value |
| --- | --- | --- | --- | --- |
| **Demographic characteristics** |  |  |  |  |
| gender (male/female) (N) | 2/8 | 5/8 | 4/8 | 0.67 |
| age (years) | 61.9±13.4 | 71.5±16.6 | 75.4 ±9.9 | **0.03** |
| **Laboratory data** |  |  |  |  |
| CRP (mg/L) | 58.8±53.1 | 81.4±54.1 | 68.0±46.4 | 0.61 |
| eGFR (ml/min/1.73m<sup>2</sup>) | 95.1±20.6 | 62.5±35.4 | 75.3±27.7 | **0.04** |
| Neutrophil count (×10<sup>3</sup> /μl) | 6.0±1.7 | 9.9±4.2 | 6.3±2.7 | **0.04** |
| Eosinophil count (×10<sup>3</sup> /μl) | 14.2±12.1 | 0.2±0.2 | 0.2±0.2 | **<0.01** |
| Platelet (×10<sup>4</sup> /μl) | 30.4±9.7 | 35.8±7.9 | 34.9±6.7 | 0.29 |
| D-dimer (μg/ml) | 2.1±0.9 | 2.0±1.2 | 1.2±0.7 | 0.06 |
| MPO-ANCA (N) | 3 | 13 | 11 | **<0.01** |
| PR3-ANCA (N) | 0 | 0 | 2 | 0.16 |
| ECP (ng/ml) | 5442±3217 | 221±173 | 340±359 | **<0.01** |
| **Disease-specific characteristics** |  |  |  |  |
| renal involvement (N) | 2 | 8 | 5 | 0.09 |
| pulmonary involvement (N) | 5 | 9 | 7 | 0.69 |
| ENT involvement (N) | 6 | 0 | 10 | **<0.01** |
| Nervous system involvement (N) | 8 | 3 | 5 | **0.02** |
| Cardiac involvement (N) | 2 | 0 | 0 | 0.08 |
| Skin lesions (N) | 7 | 2 | 1 | 0.01 |
| Clinical thrombosis (N) | 2 | 2 | 1 | 0.71 |
| BVAS | 12.2±4.2 | 12.8±7.8 | 13.9±6.1 | 0.79 |

Values given as mean ± SD or number of patients (n). Categorical variables analyzed by Fisher's exact test and continuous variables analyzed by Kruskal–Wallis test. EGPA: eosinophilic granulomatosis with polyangiitis, MPA: microscopic polyangiitis, GPA: granulomatosis with polyangiitis, CRP: C-reactive protein, eGFR: estimated glomerular filtration rate, MPO: myeloperoxidase, PR3: proteinase 3, ANCA:

### Circulating cfDNA and DNase activity in patients with AAV

We quantified circulating cfDNA by qPCR. The concentration of cf-nDNA was significantly higher in AAV patients than in HC (median [interquartile range, IQR]: EGPA 913.5 [419.8–1617.5] ng/ml, MPA 194 [83.1–357] ng/ml, GPA58.5 [33.9–158.3] ng/ml, HC 7.1 [5.7–42.2] ng/ml, respectively) (**Fig. 1a**). The concentration of cf-mtDNA was also significantly higher in EGPA and MPA patients than in HC, but there was no significant difference between GPA and HC (median [IQR]: EGPA 0.72 [0.3–1.77] pg/ml, MPA 0.18 [0.1–0.98] pg/ml, GPA 0.20 [0.03–0.43] pg/ml, HC 0.076 [0.05–0.14] pg/ml, respectively) (**Fig. 1b**). Notably, cf-nDNA levels were highest in patients with EGPA (**Fig. 1a**). Similar trends were observed in cf-mtDNA levels, but there was no significant difference between EGPA and MPA (**Fig. 1b**). The concentration of cf-nDNA was approximately a million-fold higher than that of cf-mtDNA in all groups.

**Fig. 1.**
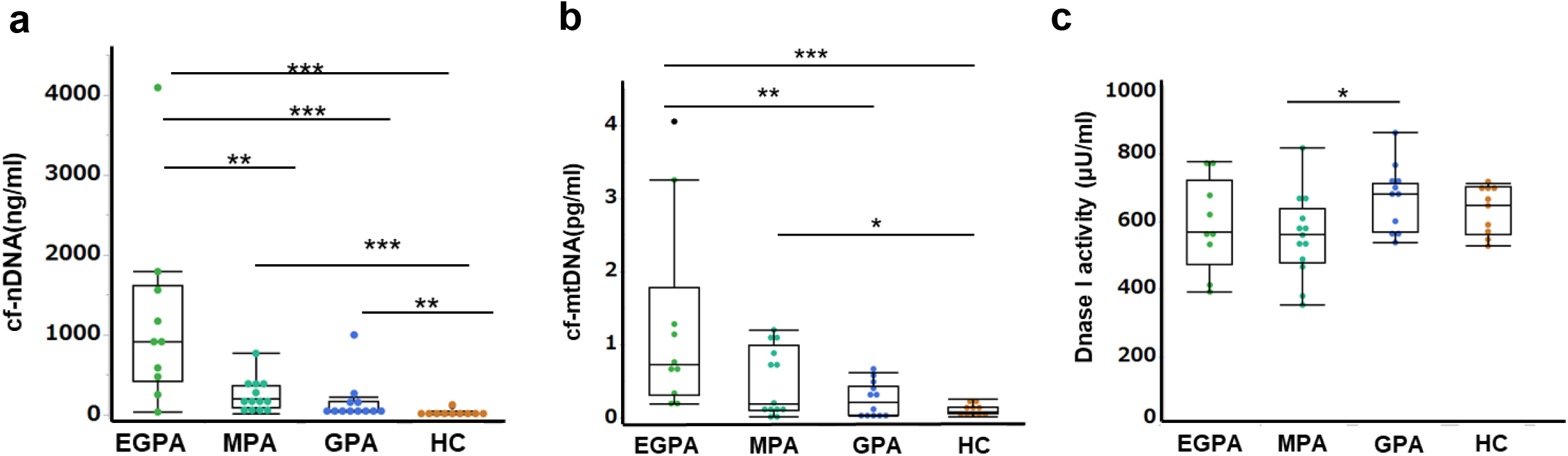
Serum concentrations of cf-nDNA and cf-mtDNA in immunosuppressive therapy-naïve patients with AAV. Concentrations of cf-nDNA in HC and patients with EGPA, MPA, and GPA before immunosuppressive therapy measured by qPCR using primer sets for the 115-bp ALU repeats. (b) Concentrations of cf-mtDNA in HC and patients with EGPA, MPA, and GPA before immunosuppressive therapy measured by qPCR using primer sets for 79-bp gene fragment of mitochondrial 16s-RNA. (c) Serum DNase1 activity before treatment measured by fluorescence spectrophotometry. One unit of DNase1 was defined as the amount of enzyme that cleaved 1.0 µmol of DNA probe per minute. EGPA: n=10, MAP: n=13, GPA: n=12, HC: n=10, \**P* < 0.05, \*\**P* < 0.01, \*\*\**P <* 0.001, ns: not significant, Mann–Whitney U test.

DNase deficiencies can lead to the persistence of DNA in the serum, which may contribute to the development of various autoimmune diseases^11^. We therefore investigated serum DNase1 activity in AAV patients in relation to the concentration of cfDNA. DNase1 activity in patients with EGPA was similar to that in the MPA, GPA, and HC groups, but activity was significantly higher in patients with GPA compared with MPA. In addition, there was no correlation between serum cf-nDNA and DNase1 activity (data not shown). These results indicated that the high levels of cfDNA in patients with EGPA were not associated with decreased serum DNase1 activity.

### Association between disease activity and cfDNA in patients with AAV

We compared the circulating cf-nDNA and cf-mtDNA concentrations before and after immunosuppressive treatment. Both cf-nDNA and cf-mtDNA levels were significantly decreased in EGPA patients after therapy (median [IQR] 291.5 [98.3–733.3] ng/ml, 0.16 [0.09–0.29] pg/ml, respectively), but levels were unchanged in patients with MPA (78.3 [29–206] ng/ml, 0.15 [0.05–0.34] pg/ml) and GPA (198.5 [58.7–537.3] ng/ml, 0.09 [0.03–0.23] pg/ml) (**Fig. 2**). These results indicated that cfDNA was only associated with disease activity in EGPA. We evaluated the association between cfDNA and BVAS, as a monitoring parameter for disease activity. The concentrations of cf-nDNA and cf-mtDNA were correlated with BVAS score in patients with EGPA, but not in patients with GPA and MPA (**Fig. 2**).

**Fig. 2.**
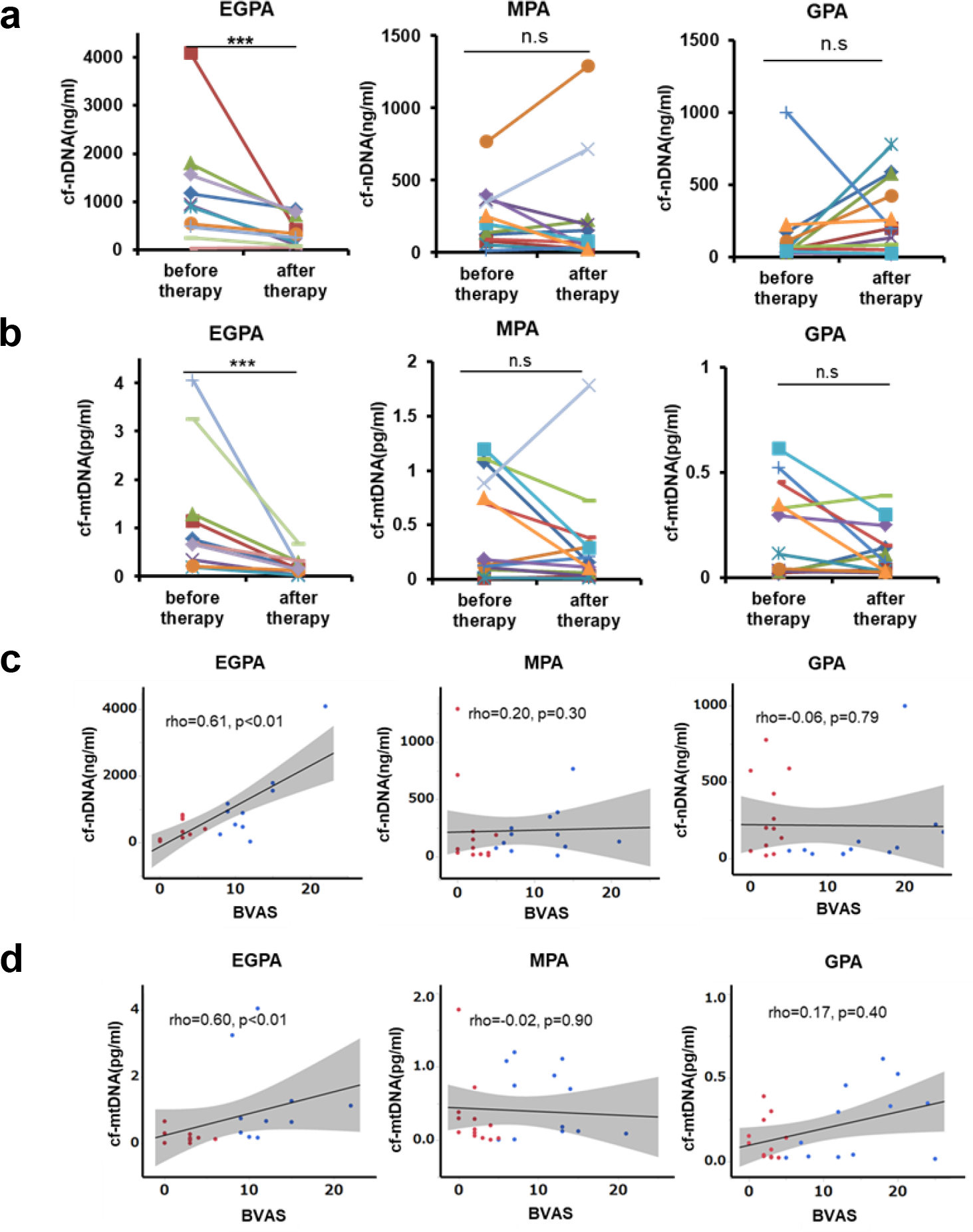
Serum cfDNA levels and disease activity in patients with AAV. (a) Concentrations of cf-nDNA and (b) cf-mtDNA in patients with EGPA, MPA, and GPA before and after immunosuppressive therapy. Same patients connected by lines. EGPA: n=10, MAP: n=13, GPA: n=12, \*\*\**P* < 0.01, ns: not significant, Wilcoxon signed rank test. (c) Correlations between serum cfDNA and disease activity in patients with AAV. A significant correlation was only observed in patients with EGPA. Blue dots: before treatment, red dots: after treatment, straight line: regression line, gray area: 95% confidence interval, Spearman’s rank correlation coefficient.

### cfDNA was associated with eosinophil count, ECP, and D-dimer level in EGPA

We also analyzed the association between cfDNA and laboratory test results in patients with AAV (**Fig. S1a**). Neutrophil count was correlated with cf-nDNA in patients with MPA and GPA, but not EGPA (**Fig. S1b-d**). However, both cf-nDNA and cf-mtDNA levels were associated with eosinophil count, ECP, and levels of the fibrin degradation product D-dimer in EGPA (**Fig. 3**). cfDNA was not correlated with D-dimer levels in the other AAVs.

**Fig. 3.**
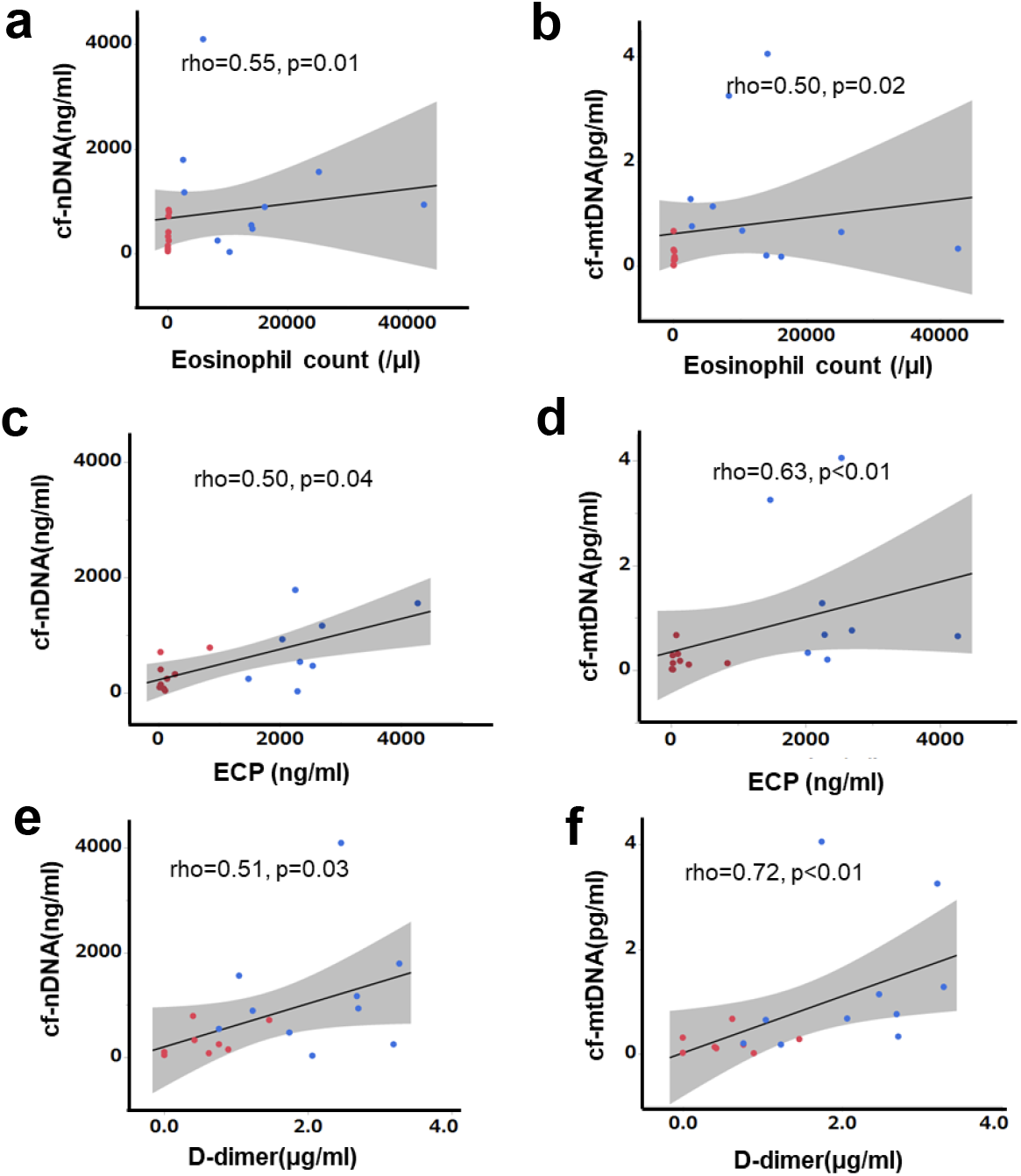
Correlations between cfDNA and eosinophil count, ECP, and D-dimer level in patients with EGPA. Eosinophil count (a, b) ECP (c,d), and D-dimer level (e, f) were associated with cf-nDNA and cf-mtDNA. Blue dots: before treatment, red dots: after treatment, straight line: regression line, gray area: 95% confidence interval, n=10, Spearman’s rank correlation coefficient.

### Presence of EETosis in small vessel thrombi in patients with EGPA

EETs have been shown to contribute to thrombus formation^33^. Given the positive correlation between peripheral blood eosinophils and D-dimer levels in patients with EGPA, we hypothesized that EETs might be associated with thrombus formation. We investigated this hypothesis in affected tissues from patients with active EGPA. Tissues were immunostained with CD31 (platelet endothelial cell adhesion molecule-1), a transmembrane glycoprotein expressed on the surface of vascular endothelial cells, and CitH3, a marker for ETs^34^. H&E staining showed massive infiltration of eosinophils and neutrophils around the small vessels, and also occluding the vessels (**Fig. 4ai**). Immunostaining of identical fields indicated several chromatolytic cells containing net-like CitH3 and DNA (**Fig. 4aii**), indicating the presence of ETs in small vessel thrombi. Staining specificities and other representative examples of CitH3-positive eosinophils in occluding vessels are shown in **Fig. S2**.

**Fig. 4.**
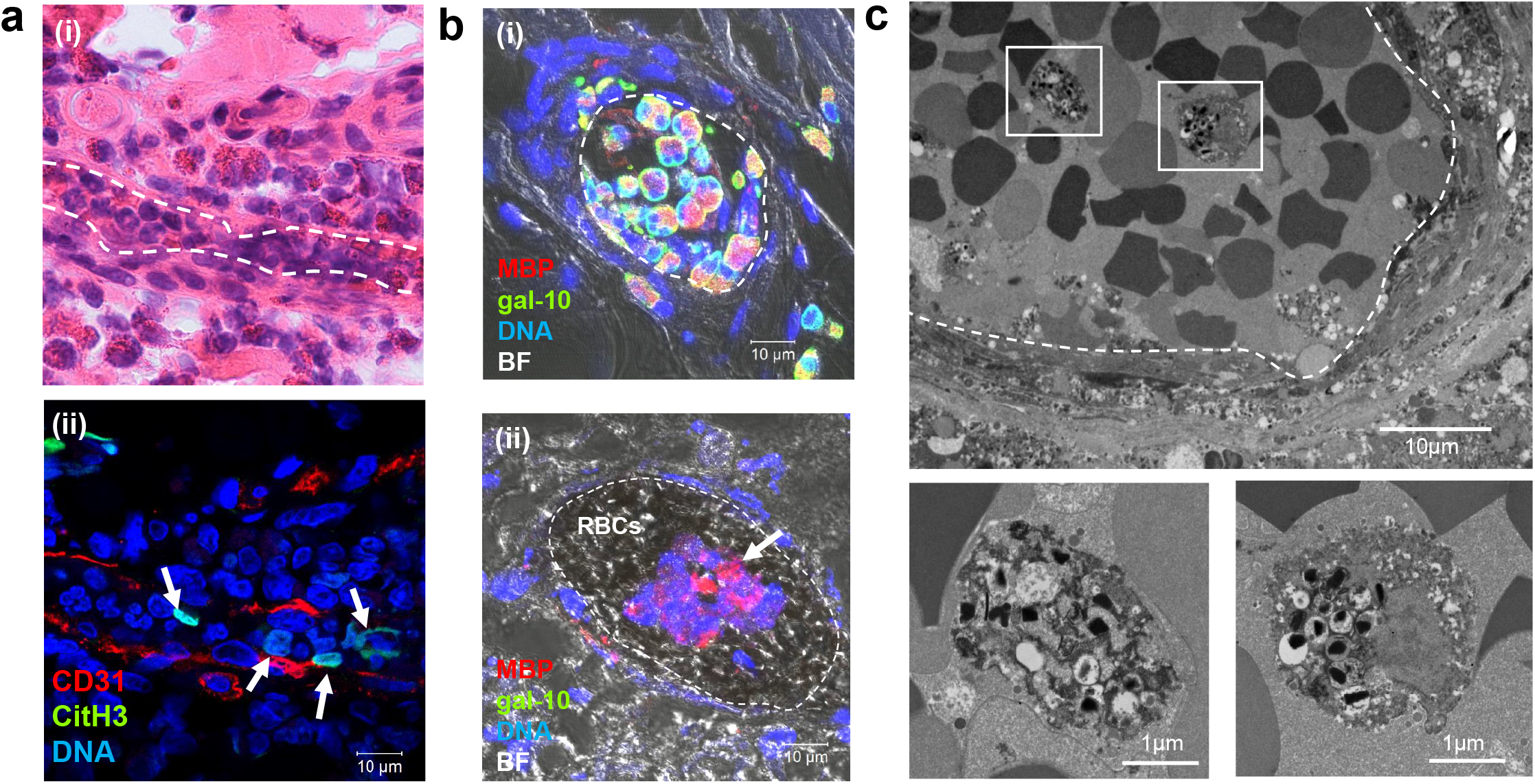
EETosis in small vessel thrombi in patients with EGPA. Inflamed skin biopsy samples from patients with EGPA were observed using H&E staining (ai) and immunostaining for CD31 and CitH3 (aii) (identical sample). Inflammatory cell infiltration including eosinophils was observed in the perivascular area and occluding small vessels (dashed line in (ai)). Net-like CitH3-positive nuclei were present in small vessels (aii, arrows). (b) Sural nerve biopsy samples were stained to detect the eosinophil granule protein MBP, eosinophil cytoplasmic protein galectin-10 (gal-10), and DNA. Merged immunofluorescence and bright field (BF) images indicated clustering of intact eosinophils in the intravascular space (bi) and clusters of EETotic eosinophils in thrombus (bii, arrow). Clustered red blood cells (RBCs) were also observed. (c) TEM of sural nerve tissues showed lytic eosinophils with EETotic morphologies in small vessel thrombus (dashed line). Boxed areas in upper panel are magnified in lower panels.

Additionally, biopsy samples were double-immunostained for the eosinophil-specific proteins, cytoplasmic protein galectin-10 and granular protein MBP, to detect intact and EETotic cells. Intact eosinophils were galectin-10- and MBP-positive (gal-10+/MBP+), whereas lytic cells with free granules were only positive for MBP (gal-10-/MBP+)^35,36^. Intravascular intact eosinophils (gal-10+/MBP+) were occasionally found clustered and adherent to the endothelium (**Fig. 4bi**). EETotic eosinophils (gal-10-/MBP+) were also observed in small vessel thrombi (**Fig. 4bii**). Staining specificities and additional representative examples are shown in **Fig. S3**.

The ultrastructural morphological characteristics of EETosis were confirmed by TEM (**Fig. 4c, Fig. S4**). Chromatolytic eosinophils with several intact granules (some showing loss of granular contents) were observed in thrombi. TEM revealed eosinophil-containing thrombi in five samples from seven patients. These results suggested that EETosis-mediated cytolytic eosinophils in thrombi might be responsible for the increases in circulating cf-nDNA and cf-mtDNA in EGPA patients.

### EETs are structurally resistant to degradation by DNase and induce platelet adhesion

EETs have previously shown bolder chromatin threads and more aggregation than NETs^21,37,38^. We further investigated the structural characteristics of EETs and NETs *in vitro,* using isolated human eosinophils and neutrophils. When EETosis and NETosis were induced in static culture conditions, there was a significant difference in spontaneous release of ETs assessed by the cell-impermeable DNA dye SYTOX (**Fig. 5a**, upper panels). The ultrastructural characteristics of NETs and EETs were also compared by SEM (**Fig. 5a**, lower panels). EETs were associated with lytic cells and did not spread spontaneously, whereas NETs were more prone to expand, showing a larger DNA area (**Fig. 5b**). EETs showed bolder chromatin threads and were more aggregated than NETs, confirming the well-conserved nucleosome structures.

**Fig. 5.**
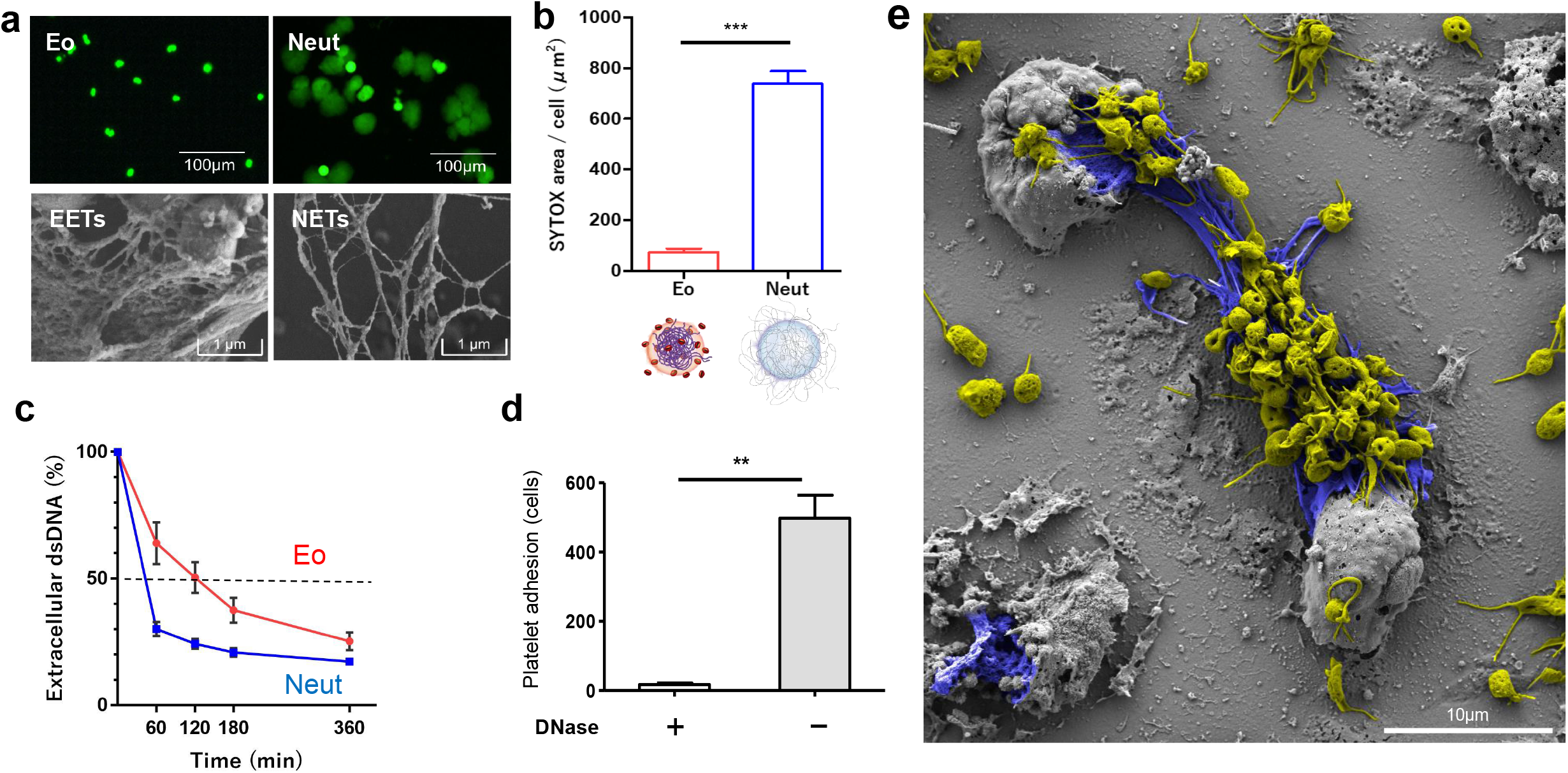
EETs are structurally stabile and induce platelet adhesion. (a, upper panels) Spontaneous spreading of EETs and NETs. Isolated eosinophils (Eo) and neutrophils (Neut) were stimulated with PMA to induce ET formation. Extracellular DNA was visualized using the cell-impermeable fluorescent DNA dye SYTOX in static culture conditions, and images were obtained using a fluorescence inverted microscope (a, lower panels). Morphological differences between EETs and NETs were determined by SEM. EETs showed bold and condensed chromatin threads. Cell-free eosinophil granules (asterisk) were also observed. (b) SYTOX area per cell was measured using ImageJ software. Results were expressed as mean ± standard deviation (SD). n = 3, \*\*\**P* < 0.001, Student’s *t*-test. (c) DNase1 was added to EETs and NETs and dsDNA was quantified using Picogreen intensity. The amount of dsDNA (DNase1/vehicle control × 100) at 0 minutes was considered as baseline. Results were expressed as mean ± SD. n = 3, \**P* < 0.05, Student’s *t*-test. (d) EETosis cells were treated with or without DNase followed by incubation with fluorescence-labeled platelet suspension. Adhered platelets per field were counted. Results were expressed as mean ± SD. n = 3, \**P* < 0.05. (e) SEM image indicated abundant platelets (yellow -colored) adhered to EETs (blue -colored). Platelets were aggregated and formed filopods. Scale bar; 10 µm.

The nucleosome structure protects DNA from fragmentation by DNase^1^. We then studied the stabilities of EETs and NETs against DNase. EETs and NETs were treated with DNase followed by measurement of dsDNA levels using the fluorescence dye Picogreen. NETs showed faster dissolution than EETs, with approximate half-lives of 42 and 120 minutes, respectively (**Fig. 5c**). Taken together, these results indicated that EETs had greater stability against exposure to DNase than NETs, likely leading to higher serum cfDNA concentrations in EGPA patients.

NETs provide a scaffold for platelet adhesion, leading to thrombus formation^39^. To test if EETs had similar properties, DNase-treated or vehicle-treated EETosis cells were incubated with fluorescence-labeled platelets suspended in plasma, and adherent platelets were quantified. The platelets adhered to the EETosis cells, and adherence was decreased when EETs were removed by DNase (**Fig. 5d**). SEM showed abundant platelets adhered to EETs and EETotic cells (**Fig. 5e**). Filopod formation and aggregation of platelets indicated their active state.

## Discussion

AAV is a life-threatening disease with complex management, due to the lack of suitable biomarkers. Elevated or persistently positive ANCA titers as an indicator of disease activity or relapse in patients with AAV remains controversial^40,41^, and commonly measured laboratory markers, such as absolute eosinophil count, serum IgE, erythrocyte sedimentation rate, and C-reactive protein, have limitations as biomarkers of disease activity or predictors of flare in patients with EGPA^42^.

Although a previous report showed increased levels of cf-nDNA in patients with EGPA, the study only included patients in remission^24^. The current study indicated that cfDNA, including nDNA and mtDNA, might be useful biomarkers reflecting disease activity in EGPA. Compared with HC, patients in all AAV groups showed elevated serum levels of cfDNA, while both cf-nDNA and cf-mtDNA levels were higher in patients with EGPA compared with those with GPA or MPA. cfDNA levels decreased in response to immunosuppressive therapy and were also correlated with disease activity assessed by BVAS, but only in patients with EGPA. A previous study showed that serum cf-mtDNA levels were higher in patients with active GPA compared with those in remission^23^. The apparent discrepancy between these results could be due to differences in patient backgrounds, study periods, and treatment regimens between the studies.

Serum cfDNA was positively correlated with eosinophil count but not with neutrophil count in patients with EGPA, while positive correlations between neutrophil count and cf-nDNA were observed in patients with GPA and MPA. Eosinophils and neutrophils are major inflammatory cells in the pathogenesis of AAVs, and it is therefore conceivable that these cells may contribute to the increased serum cfDNA levels in patients with AAV.

Eosinophilia is thought to increase the risk of thrombosis^43^. The frequency of thrombosis in patients with EGPA was reviewed by Ames et al., who found prevalences of arterial and venous occlusion of 3.1%–18.7% and 5.8%–30%, respectively^44^. Kanno et al. reported that elevated D-dimer levels (>2.5 μg/ml) were associated with concomitant systemic thrombotic symptoms in EGPA^45^. Several mechanisms have been proposed to account for the procoagulant effects of eosinophil-induced factors. For instance, eosinophils store and release tissue factor^46^, and cationic proteins in eosinophil granules can act as platelet agonists to increase vascular permeability, stimulate the activation of factor XII, and inhibit heparin, leading to thrombosis^44^. Interestingly, a recent histological study indicated that 0.2%–3% of vessels in nerve tissues from patients with EGPA were occluded by intraluminal eosinophils^47^.

Immunothrombosis has been an active area of research as a process whereby NETs/NETosis play a beneficial role in host defense by trapping and killing pathogens at the expense of promoting thrombosis ^48^. In turn, unregulated immunothrombosis may lead to thrombotic disorders, including venous thromboembolism^48^. The importance of EETs/EETosis in the pathogenesis of thrombosis has not been well understood. Here, we provided direct evidence of EETs/EETosis within the thrombus in patients with EGPA. EETosis comprises active cytolysis through dissolution of the nuclear and plasma membranes, leading to total cell degranulation and the release of net-like chromatin structures. Eosinophils are thought to degranulate in the tissue, but not in the blood vessels^49^. However, an *ex vivo* study demonstrated cytolytic eosinophil degranulation in coagulated blood^50^. Mukerjee et al. showed that sputum ANCA from patients with EGPA was capable of inducing EETs from eosinophils *in vitro*^51^. In addition, immobilized immunoglobulin or platelet activating factor with interleukin (IL)-5 were shown to induce EETosis^38^. It is therefore possible that eosinophils are activated to undergo EETosis in response to local stimuli produced in the thrombus microenvironment (**Fig. S5**). Given that extracellular histones were shown to be toxic in vascular endothelial cells^52^ and to promote coagulation^48^, EETs together with granular proteins and other damage-associated molecular patterns might damage endothelial cells, further promoting vascular thrombosis. Indeed, EETs have been shown to promote atherosclerotic plaque formation and thrombosis^33^.

Among the three types of AAVs, cf-nDNA levels were markedly higher in patients with EGPA, despite comparable serum DNase levels. Previous studies indicated that NETs were composed of stacked cylindrical nucleosomes consisting of 5-10 nm smooth stretches and 25–50 nm globular domains^37^. In contrast, EETs consisted mostly of fibers with a diameter of 25–35 nm^21,38^. Our results confirmed that EETs had well-conserved nucleosome structures with bolder chromatin threads and were more aggregated than NETs. EETs had an approximately four-fold longer half-life than NETs in the presence of DNase, probably because of the protective effect of the intact nucleosome structure. Our current data highlighted the greater stability of EETs, likely leading to the higher serum cfDNA concentrations in EGPA patients. The cf-mtDNA concentration in patients with EGPA was approximately a million times lower than the cf-nDNA concentration. The low levels of mtDNA in human eosinophils^53^ and its instability due to the lack of nucleosome structure might explain this difference. This study was limited by the small number of patients with EGPA and the difficulty in providing direct evidence for the association between serum cfDNA and EETs/EETosis in thrombi. Unfortunately, there are currently no assays for distinguishing between EETosis-derived and non-eosinophil-derived cfDNA. Further studies are therefore needed to understand the cell origin, production, and clearance of cfDNA in patients with EGPA.

In summary, this study provides the first evidence for an association between serum cfDNA levels and disease activity in patients with EGPA. Increased cfDNA levels appear to be correlated with eosinophil count, ECP, and D-dimer level, likely due to the presence of EETs/EETosis within the thrombus in patients with EGPA. The stability of EETs against DNase suggest the pathophysiological importance of eosinophils. These results shed light on the possible suitability of serum cfDNA as a biomarker reflecting EETs/EETosis and thrombus formation in patients with EGPA. Eosinophil-targeting therapy using anti-IL-5 antibody is currently clinically available for EGPA^54^. Further investigations are needed to understand the pathological roles of eosinophils in EGPA.

## Data Availability

The data that support the findings of this study are available from the corresponding author, [T.H.], upon reasonable request.

## Acknowledgments

The authors are grateful to Noriko Tan and Sachie Kitano for outstanding technical assistance, and to Satomi Misawa for outstanding assistance with drawing the figure. We also thank Susan Furness, PhD, from Edanz Group (https://en-author-services.edanz.com/ac) for editing a draft of this manuscript. This study was funded in part by a Research Grant on Allergic Disease and Immunology from the Japan Agency for Medical Research and Development (JP20ek0410055 to SU), the Mochida Memorial Foundation for Medical and Pharmaceutical Research, AstraZeneca Evidence Connect Externally Sponsored Research (SU), the Japanese Society of Laboratory Medicine Fund for Promotion of Scientific Research (SU), JSPS KAKENHI (20K08794 to SU; 19K17898 to YK). This work was also supported by a Grant-in-Aid for Researchers, Hyogo College of Medicine, 2019 (TH).

## Authorship contributions

Teppei Hashimoto and Shigeharu Ueki designed and performed experiments and wrote the manuscript. Yosuke Kamide, Nobuyuki Oka, Hiroki Takeuchi, and Kyoko Kanno provided clinical samples and edited the manuscript. Yui Miyabe, and Mineyo Fukuchi performed *in vitro* and immunostaining experiments. Yuichi Yokoyama, Tetsuya Furukawa, and Naoto Azuma provided clinical information and edited the manuscript. Akemi Ishida-Yamamoto, Masami Taniguchi, and Akira Hashiramoto contributed scientific advice and edited the manuscript. Kiyoshi Matsui supervised the research and edited the manuscript.

