## Supplemental information for "Increased circulating cell-free DNA in eosinophilic granulomatosis with polyangiitis: implications for eosinophil extracellular traps and immunothrombosis"

### Supporting Materials and Methods

#### Real-time PCR for quantification of cfDNA

Real-time PCR for cf-nDNA quantification was carried out in a reaction volume of 20 μl containing 0.1 μl of the extracted cf-nDNA sample, 0.8 μl (0.2 μM) of each primer, 8.3 μl of distilled water, and 10 μl of SYBR Green I Master Mix (Roche Diagnostics). Real-time PCR was performed as follows: denaturation for 30 seconds at 95°C, annealing for 30 seconds at 60°C, and extension/synthesis for 30 seconds at 72°C for 35 cycles, using a LightCycler 480 Instrument II (Roche Diagnostics). Real-time PCR for cf-mtDNA quantification was also carried out in a reaction volume of 20 μl containing 1 μl of the extracted cf-mtDNA sample, 0.8 μl (0.4 μM) of each primer, 7.4 μl of distilled water, and 10 μl of SYBR Green I Master Mix. Real-time PCR was performed as follows: denaturation for 15 seconds at 95°C, primer annealing for 60 seconds, and extension/synthesis at 60°C for 40 cycles, using a LightCycler 480 Instrument II. A standard curve was created using a 490-bp DNA fragment, as described previously[27].

#### Measurement of DNase1 activity

DNase1 activity in serum was measured using a DNase1 Activity Assay Kit (Bio Vision, CA, USA). Briefly, enzyme activity was detected by cleavage of a DNA probe to produce a fluorescent DNA product, which was then measured at Ex/Em=651/681 nm in kinetic mode every 30 seconds for 90 minutes at 37°C, using an Infinite M200 PRO plate reader (Tecan, Zurich, Switzerland).

#### Measurement of serum ECP

Serum ECP concentrations in patients with AAV were measured using a Human ECP enzyme-linked immunosorbent assay kit (Aviscera Bioscience, Inc., CA, USA).

**TEM**

Ultrathin sections (80 nm) were stained with uranyl acetate and lead citrate, mounted on uncoated 200-mesh copper grids (Ted Pella, Redding, CA, USA), stained for 20 minutes in 4% uranyl acetate and 0.5% lead citrate, and viewed under a transmission electron microscope (H-7650, Hitachi, Tokyo, Japan) at 100 kV.

**SEM**

Purified eosinophils and neutrophils ($2 \times 10^5$ cells in 0.3% BSA RPMI) were stimulated with 10 ng/ml PMA for 180 minutes on glass slides and then prepared for SEM observation. Briefly, samples were fixed with 1.25% glutaraldehyde/1% paraformaldehyde, dehydrated in a graded series of ethanols, and reduced to t-butyl alcohol. They were then freeze-dried, coated with osmium using an osmium coater (NEOC, Meiwafosis, Tokyo, Japan), and observed on a field emission scanning electron microscope (SU-8020; Hitachi High-Technologies, Tokyo, Japan or JSM-7800F; JEOL, Tokyo, Japan).

### Supporting data

#### Fig.S1

a

| EGPA | Abs WBC | Abs Neut | Abs Eos | Plt | CRP | eGFR | D-Dimer | ECP | IgE |
|---|---|---|---|---|---|---|---|---|---|
| cfnDNA | 0.32 (0.16) | -0.04 (0.84) | **0.55 (0.01)** | **0.66 (<0.01)** | 0.42 (0.06) | 0.08 (0.71) | **0.51 (0.03)** | **0.50 (0.04)** | 0.23 (0.38) |
| cfmtDNA | 0.41 (0.07) | 0.04 (0.86) | **0.50 (0.02)** | 0.37 (0.1) | **0.51 (0.02)** | -0.10 (0.64) | **0.72 (<0.01)** | **0.63 (<0.01)** | 0.47 (0.09) |

| MPA | Abs WBC | Abs Neut | Abs Eos | Plt | CRP | eGFR | D-Dimer | ECP |
|---|---|---|---|---|---|---|---|---|
| cfnDNA | **0.48 (0.01)** | **0.43 (0.02)** | 0.23 (0.26) | **0.4 (0.04)** | **0.46 (0.02)** | **0.42 (0.03)** | -0.15 (0.56) | **0.67 (<0.01)** |
| cfmtDNA | -0.02 (0.88) | -0.001 (0.99) | 0.1 (0.62) | 0.31 (0.13) | 0.3 (0.12) | 0.32 (0.1) | 0.39 (0.13) | **0.63 (<0.01)** |

| GPA | Abs WBC | Abs Neut | Abs Eos | Plt | CRP | eGFR | D-Dimer | ECP |
|---|---|---|---|---|---|---|---|---|
| cfnDNA | **0.42 (0.04)** | **0.6 (<0.01)** | **-0.46 (0.02)** | -0.15 (0.47) | -0.17 (0.4) | -0.03 (0.88) | 0.11 (0.65) | 0.28 (0.21) |
| cfmtDNA | -0.001 (0.99) | -0.12 (0.55) | 0.23 (0.27) | 0.31 (0.13) | 0.29 (0.16) | 0.24 (0.26) | -0.46 (0.06) | 0.31 (0.15) |

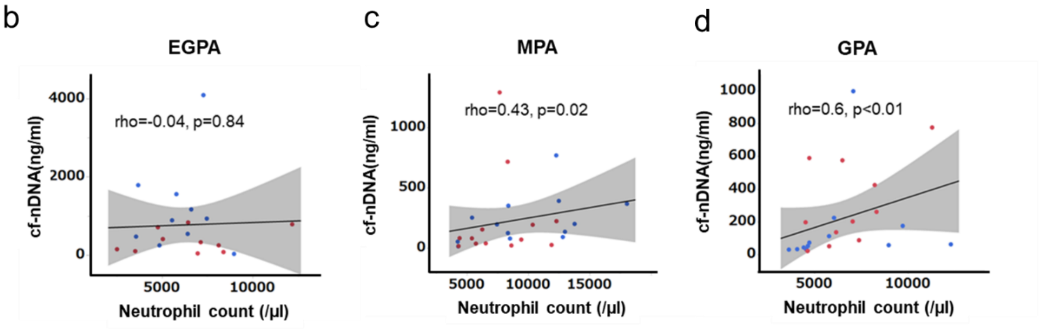


**Fig. S1** Correlations between cfDNA levels and laboratory test results in patients with AAV.

(a) Spearman's correlation coefficients presented with *P*-values in parentheses. Areas highlighted in gray indicate significant correlations. WBC: white blood cell count, Neut: neutrophil count, Eos: eosinophil count, ECP: eosinophilic cationic protein. Dot-plots of correlations between neutrophil count and serum cf-nDNA levels in patients with EGPA (b), MPA (c), and GPA (d). Blue dots: before treatment, red dots: after treatment, straight line: regression line, gray area: 95% confidence interval, EGPA: n=10, MAP: n=13, GPA: n=12, Spearman's rank correlation coefficient.

**Fig.S2**

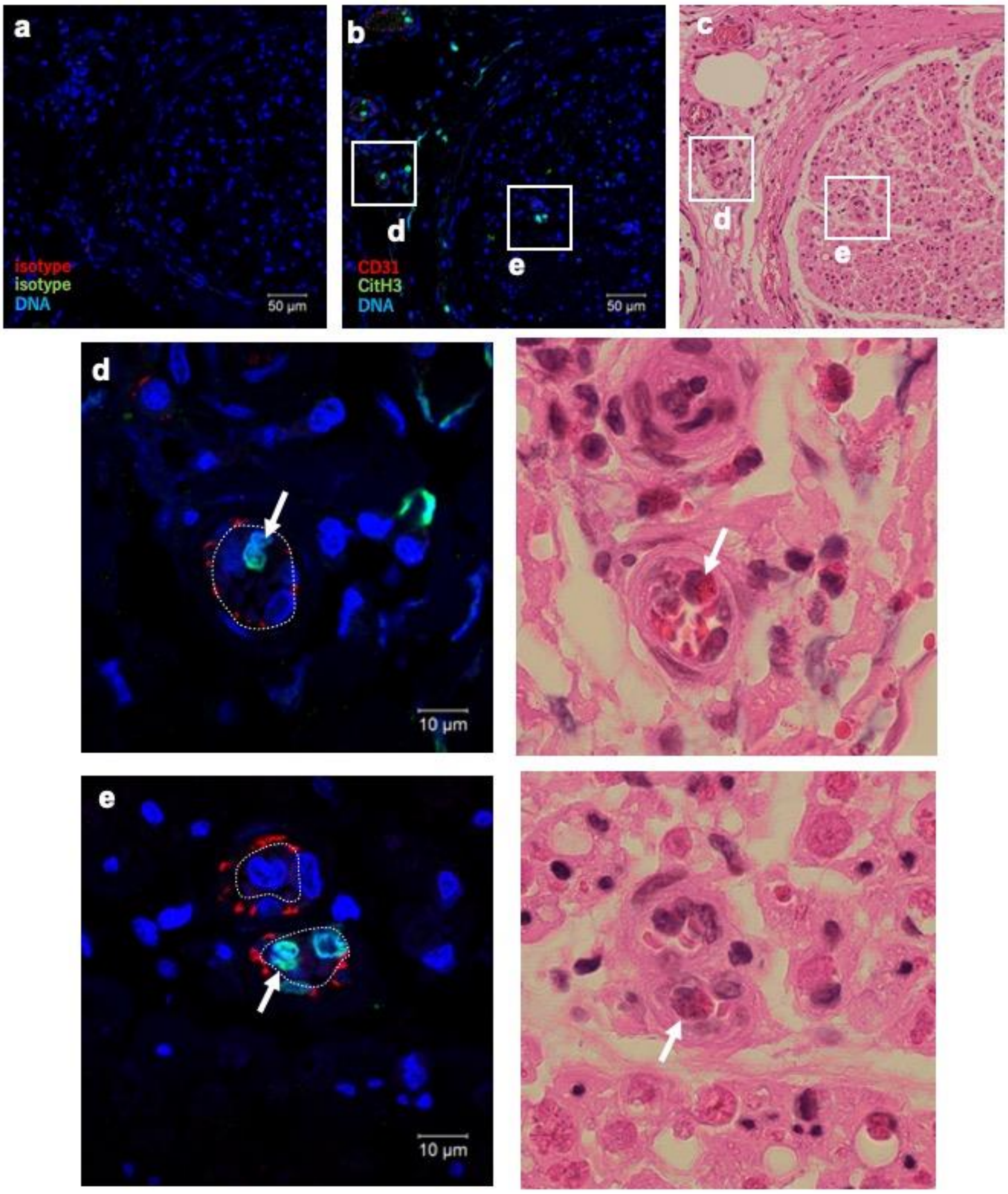

a
isotype
isotype
DNA
50 µm
b
d
e
CD31
CitH3
DNA
50 µm
c
d
e
d
10 µm
e
10 µm

**Fig.S2 (cont'd)**

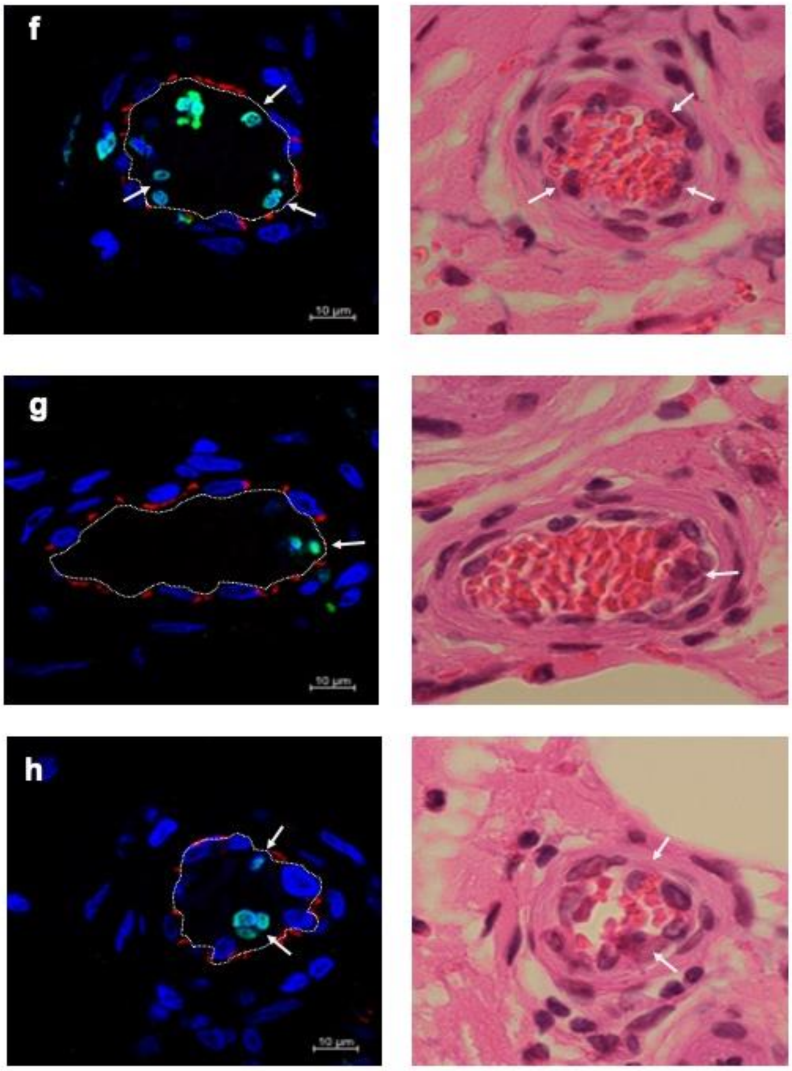


**Fig. S2** Citrullinated histone-positive eosinophils in blood vessel thrombi.

Sural nerve biopsy samples from patients with EGPA were assessed by immunostaining with (a) isotype-matched control antibodies and (b) specific antibodies for CD31 (red) and CitH3 (green), and (c) by H&E staining. Images (b) and (c) show identical fields from same sample section; (a) shows serial section from same sample. Images (a) and (b) obtained at same confocal microscopy setting, indicating staining specificities. (c) Magnified image of nerve trunk. (d) Extrinsic vessel and (e) intrinsic vessel, both occluded by inflammatory cells and red blood cells. (f-h) Additional representative images of thrombi from different donors. Arrows indicate CitH3-stained chromatolytic eosinophils.

Fig.S3

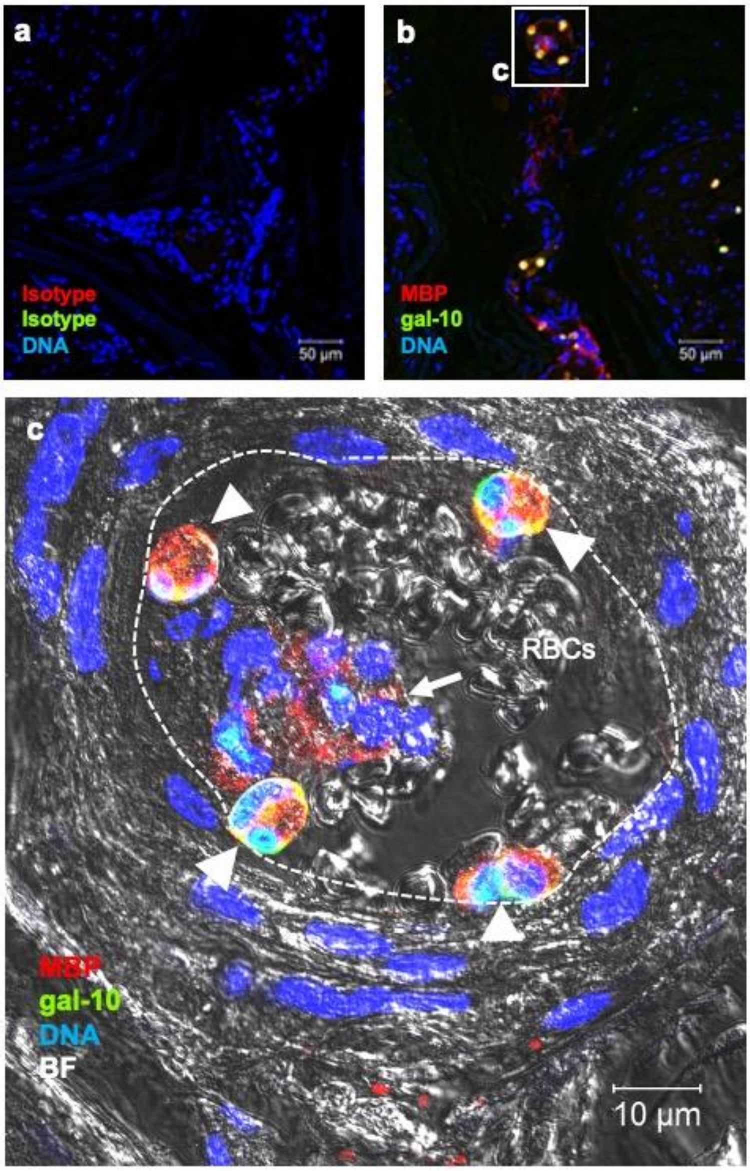
a
Isotype
Isotype
DNA
50 µm
b
c
MBP
gal-10
DNA
50 µm
c
RBCs
MBP
gal-10
DNA
BF
10 µm

**Fig.S3 (cont'd)**

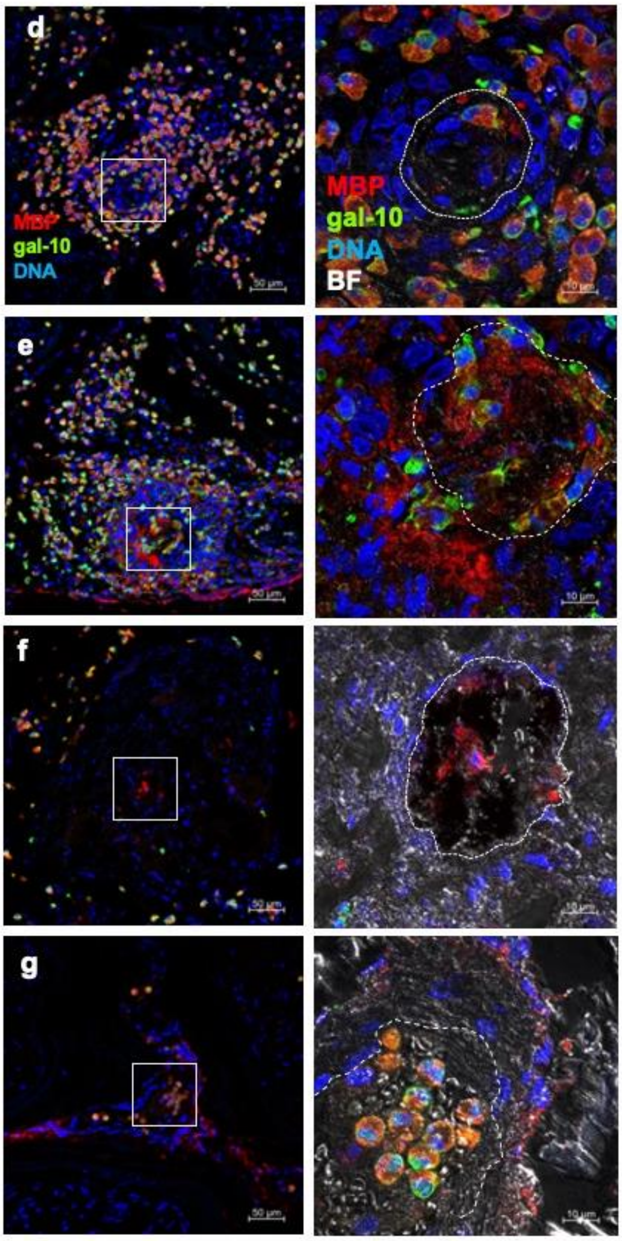


**Fig. S3** Intact and lytic eosinophils in small vessel thrombi.

Sural nerve biopsy samples from patients with EGPA were assessed by immunostaining using (a) isotype-matched control antibodies and (b) specific antibodies for MBP (red) and galectin-10 (green). Images (a) and (b) obtained at same confocal microscopy setting, indicating staining specificities. (c) Magnified image of extrinsic vessel showing intravascular thrombus with red blood cells (RBCs), intact eosinophils (arrowheads), and clustered lytic eosinophils (arrow). (d-g) Additional representative images. The cytolytic eosinophils in thrombi were present in various degree.

Fig.S4

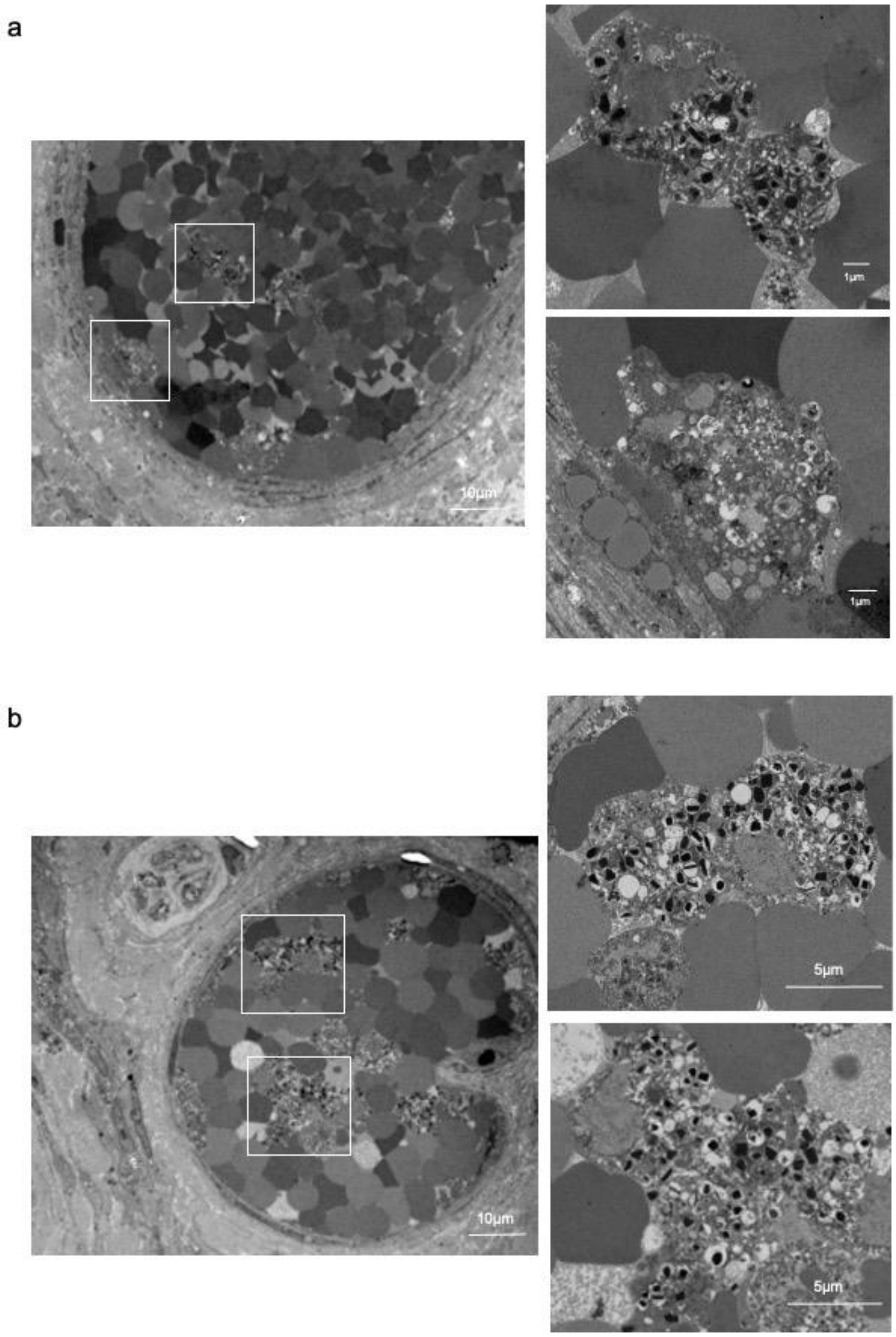


**Fig. S4** TEM of blood vessel thrombi in patients with EGPA.

(a, b) Additional representative TEM images are shown. Thrombus in sural nerve tissues showing lytic eosinophils with EETotic morphologies. Boxed areas in left panels are magnified in right panels.

**Fig.S5**

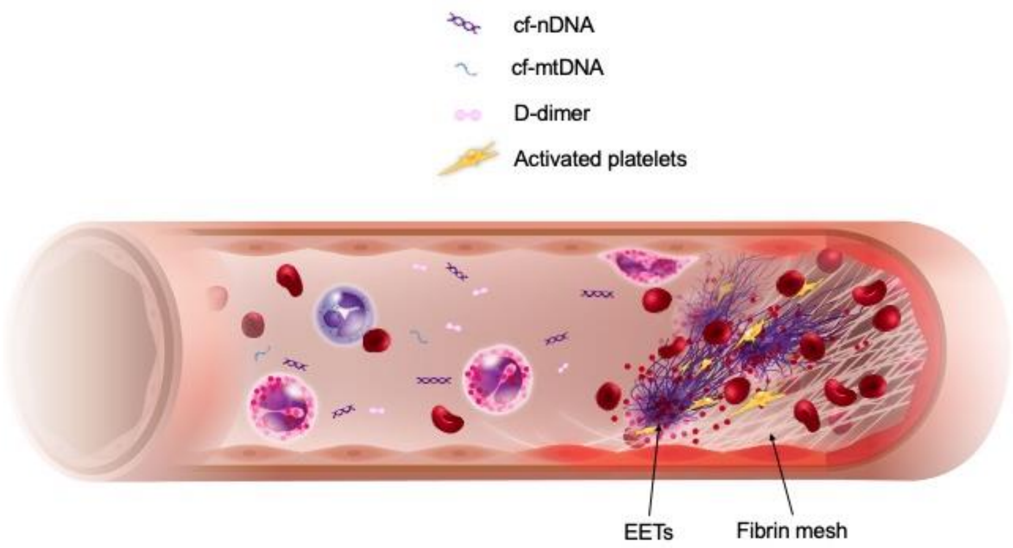


**Fig. S5** EETs/EETosis in thrombus in EGPA.

Graphic diagram showing thrombus in affected tissue in EGPA. Activated eosinophils can undergo programmed cell death via EETosis to release condensed chromatin structure EETs. EETs might provide a scaffold for platelets and promote vascular injury, leading to immunothrombosis. Increased cf-nDNA and cf-mtDNA levels might originate from cytolytic eosinophils in the thrombus.

### Supporting video legends

**Supporting Video 1.** EETosis mediates cell death and release of nuclear-derived extracellular DNA.

To ascertain if the EETs were mainly derived from the nucleus, isolated eosinophils were stained with the cell-permeable DNA dye Hoechst 33342 and suspended in 0.3% BSA/RPMI medium containing the cell-impermeable DNA dye SYTOX green. The cell suspension was seeded in a glass chamber and stimulated with 10 ng/ml PMA at 37°C in a $CO_2$ incubator. Bright-field and fluorescence images were obtained every 5 minutes using a BZ-X800 fluorescence microscope (Keyence, Tokyo, Japan). The nuclei became round and stained green, indicating cell death. Note the absence of catapult-like release of filamentous extracellular DNA.

Table S1 Patient characteristics after immunosuppressive therapy

| | EGPA (n=10) | MPA(n=13) | GPA(n=12) | *p* value |
|---|---|---|---|---|
| **Medication** | | | | |
| Initial GC dose (mg) | 44.0±7.0 | 40.8±12.6 | 39.6±6.9 | 0.55 |
| methylprednisolone pulse (N) | 7 | 5 | 6 | 0.58 |
| intravenous CY (N) | 0 | 2 | 0 | 0.32 |
| rituximab (N) | 1 | 5 | 6 | 0.07 |
| azatioprine (N) | 2 | 3 | 3 | 0.96 |
| therapy duration (days) | 61.6±20 | 49.3±19.5 | 57.4±22.5 | 0.25 |
| **Laboratory data after therapy** | | | | |
| CRP (mg/L) | 3.9±8.2 | 1.9±2.2 | 1.6±1.9 | 0.89 |
| eGFR (ml/min/1.73m²) | 111.8±26.2 | 61.1±26.7 | 72.5±21.4 | **<0.01** |
| Neutrophil count (×$10^3$/µl) | 6.5±2.8 | 7.8±2.7 | 6.4±2.0 | 0.40 |
| Eosinophil count (×$10^3$/µl) | 0.07±0.07 | 0.04±0.07 | 0.02±0.03 | 0.22 |
| Platelet (×$10^4$/µl) | 22.7±7.1 | 22.8±5.2 | 22.1±5.4 | 0.39 |
| D-dimer (µg/ml) | 0.6±0.5 | 1.6±1.6 | 1.5±1.7 | 0.96 |
| ECP (ng/ml) | 170±263 | 31.7±13.8 | 71.2±76.4 | 0.15 |
| **Disease-specific characteristics after therapy** | | | | |
| renal involvement (N) | 1 | 1 | 2 | 1.0 |
| pulmonary involvement (N) | 0 | 0 | 5 | **<0.01** |
| ENT involvement (N) | 1 | 6 | 0 | **<0.01** |
| Nervous system involvement (N) | 7 | 3 | 3 | 0.052 |
| Cardiac involvement (N) | 0 | 0 | 0 | 1.0 |
| Skin lesions (N) | 0 | 0 | 0 | 1.0 |
| Clinical thrombosis (N) | 2 | 2 | 1 | 0.71 |
| BVAS | 2.5±1.9 | 2±1.7 | 2.3±1.5 | 0.66 |

Values given as mean ± SD or number of patients (n). Categorical variables analyzed by Fisher's exact test and continuous variables analyzed by Kruskal–Wallis test. EGPA: eosinophilic granulomatosis with polyangiitis, MPA: microscopic polyangiitis, GPA: granulomatosis with polyangiitis, CRP: C-reactive protein, eGFR: estimated glomerular filtration rate, BVAS: Birmingham Vasculitis Activity Score, ENT: ear, nose, and throat, GC: glucocorticoid, CY: cyclophosphamide
